# Integrating vaccination with short-term behavioral guidance enables mpox outbreak control

**DOI:** 10.64898/2026.05.26.26354088

**Authors:** Davide Maniscalco, Olivier Robineau, Pierre-Yves Boëlle, Alexandra Mailles, Harold Noël, Arnaud Tarantola, Annie Velter, Vittoria Colizza

**Affiliations:** Sorbonne Université, INSERM, Institut Pierre Louis d’Épidémiologie et de Santé Publique, IPLESP, Paris, France; Université Lille, Centre Hospitalier de Tourcoing, ULR 2694-METRICS: Évaluation des technologies de santé et des pratiques médicales, Lille, France; Santé publique France, Saint Maurice, France; Santé publique France Regional Office, Saint-Denis, Île-de-France, France; Aix Marseille Univ, Inserm, IRD, SESSTIM, Sciences Economiques & Sociales de la Santé & Traitement de l’Information Médicale, ISSPAM, Marseille, France

## Abstract

**Background:** Despite the decline of the 2022 global outbreak, mpox remains an ongoing public health concern, with persistent transmission and emerging viral clades sustaining resurgence risk. Improving preparedness and response is a priority, yet it remains unclear how best pre-exposure vaccination and community response can effectively limit transmission under realistic conditions and whether behavioral adaptation is critical.

**Methods:** We used a stochastic data-driven Susceptible-Exposed-Infected-Isolated-Recovered agent-based network model of mpox transmission among men who have sex with men in the Paris region, parameterized with sexual behavioral data and calibrated to surveillance data from the 2022 outbreak. We evaluated counterfactual scenarios by varying vaccination timing, rollout speed, prioritization, and behavioral responses.

**Results:** Here we show that, with respect to the 2022 epidemic in the Paris region, vaccination alone delivered at the observed rollout speed would not have reproduced the observed epidemic decline, even if initiated the day of the first European alert, corresponding to 12 days before the first case was reported in France. Under our modelling assumptions, reproducing the empirical curve through vaccination alone would have required more than a fourfold increase in rollout speed. Large-scale and long-term reductions in sexual contacts remain instrumental to limit the epidemic size, although earlier vaccination reduces the proportion of MSM needing to change behavior. In contrast, short-term behavioral measures adopted by the vaccinees, such as sexual abstinence during the 14-day immunity-building period, combined with moderately faster vaccine rollout could reproduce the observed curve without large-scale behavioral adaptations (+68% rollout speed relative to the observed rate with 50% compliance to short-term sexual abstinence; +34% with 75% compliance). Targeting individuals with higher sexual activity further improved intervention efficiency.

**Conclusions:** Under realistic reactive vaccination scenarios, mpox control still requires strong behavioral responses. Combining timely vaccination with short-term behavioral change guidance at vaccine administration offers a feasible path to limit transmission and strengthen outbreak preparedness and response.

## Introduction

Mpox remains an ongoing public health concern despite the decline of the 2022 global outbreak^1,2^. Following the spread of subclade IIb across 108 countries with over 84,000 reported cases^3^, transmission has persisted at low levels^1^. The recent emergence of Clade Ib cases^4,5^ in Europe without travel history^6^ underscores the continued risk of resurgence. Although the overall risk for the general population remains low, it is moderate among men who have sex with men (MSM*)^6^ and mpox continues to pose a significant burden in parts of Africa^2^. In this context, improving preparedness for future outbreaks is a key public health priority.

The decline of the 2022 outbreak was not driven by a single mechanism and varied across settings^7–13^, reflecting differences in the timing and scale of interventions. In several countries, vaccination campaigns were implemented after the epidemic peak. The downturn was largely attributed to behavioral changes^7,9,14,8,15^, infection-induced immunity^12,16^, or self-isolation^17^. In contrast, where vaccination was deployed earlier, it played a more substantial role in reducing transmission^10^. These observations highlight the critical importance of intervention timing^15^, but leave the question open of how vaccination and behavioral responses interact to abate an epidemic.

The Paris region provides a well-characterized example of this interplay. Modeling and survey data attributed the outbreak decline primarily to large-scale behavioral adaptations among MSM^7^, as the pre-exposure vaccination campaign began after the epidemic peak and therefore did not contribute to the observed downturn. Although vaccine uptake was substantial among groups at risk (especially HIV-PrEP users) once available^18,19^, early access was limited and largely restricted to post-exposure prophylaxis, reaching only a small fraction of the population^7,20^. This setting offers a unique opportunity to assess how earlier and more intensive vaccination could have altered the course of the epidemic.

This raises a central question: under what conditions could vaccination have reduced or replaced the large-scale behavioral adaptations observed in 2022? More specifically, how early and how fast must a vaccination campaign be deployed to meaningfully limit transmission during an explosive outbreak? To address these questions, we used the 2022 Paris epidemic as a reference and developed a data-driven network model of mpox transmission to evaluate counterfactual vaccination strategies. We quantified how vaccination timing and rollout interact with behavioral adaptation (large-scale or localized), and assessed whether earlier and faster vaccination could have reproduced the observed epidemic decline or reduced the magnitude of community response required.

We show that vaccination alone at the observed rollout speed is insufficient to reproduce the rapid decline of the 2022 epidemic, even when initiated substantially earlier, and that large-scale behavioral adaptation remains necessary under these conditions. Achieving a comparable epidemic trajectory through vaccination alone requires substantially faster rollout. Combining earlier vaccination with short-term post-vaccination sexual abstinence reduces this effort without requiring large-scale behavioral adaptation, with further gains from prioritizing individuals with higher sexual activity. These findings highlight the importance of integrating timely vaccination with feasible, targeted behavioral guidance in preparedness for future mpox outbreaks.

## Methods

To quantify how vaccination preparedness and response - including stocks availabilities, efficient distribution and uptake – could have altered the course of the 2022 mpox outbreak in the Paris region, we used the observed epidemic trajectory (**Figure 1a**) as a benchmark and evaluated counterfactual vaccination strategies. Specifically, we asked whether earlier initiation and faster rollout of pre-exposure vaccination could have reproduced the observed epidemic decline without large-scale behavioral adaptation, or reduced the magnitude of community response required. Using a data-driven network model of sexual transmission among MSM, we examined four preparedness scenarios combining different assumptions on vaccination timing, rollout speed, and distinct behavioral responses, including community-wide reductions in sexual contacts and short-term post-vaccination abstinence. These scenarios progressively explore whether vaccination alone could reproduce the observed downturn, how much behavioral change would still be required despite earlier vaccination and what level of acceleration would have been necessary for vaccination to substantially reduce the need for spontaneous risk reduction.

**Figure 1.**
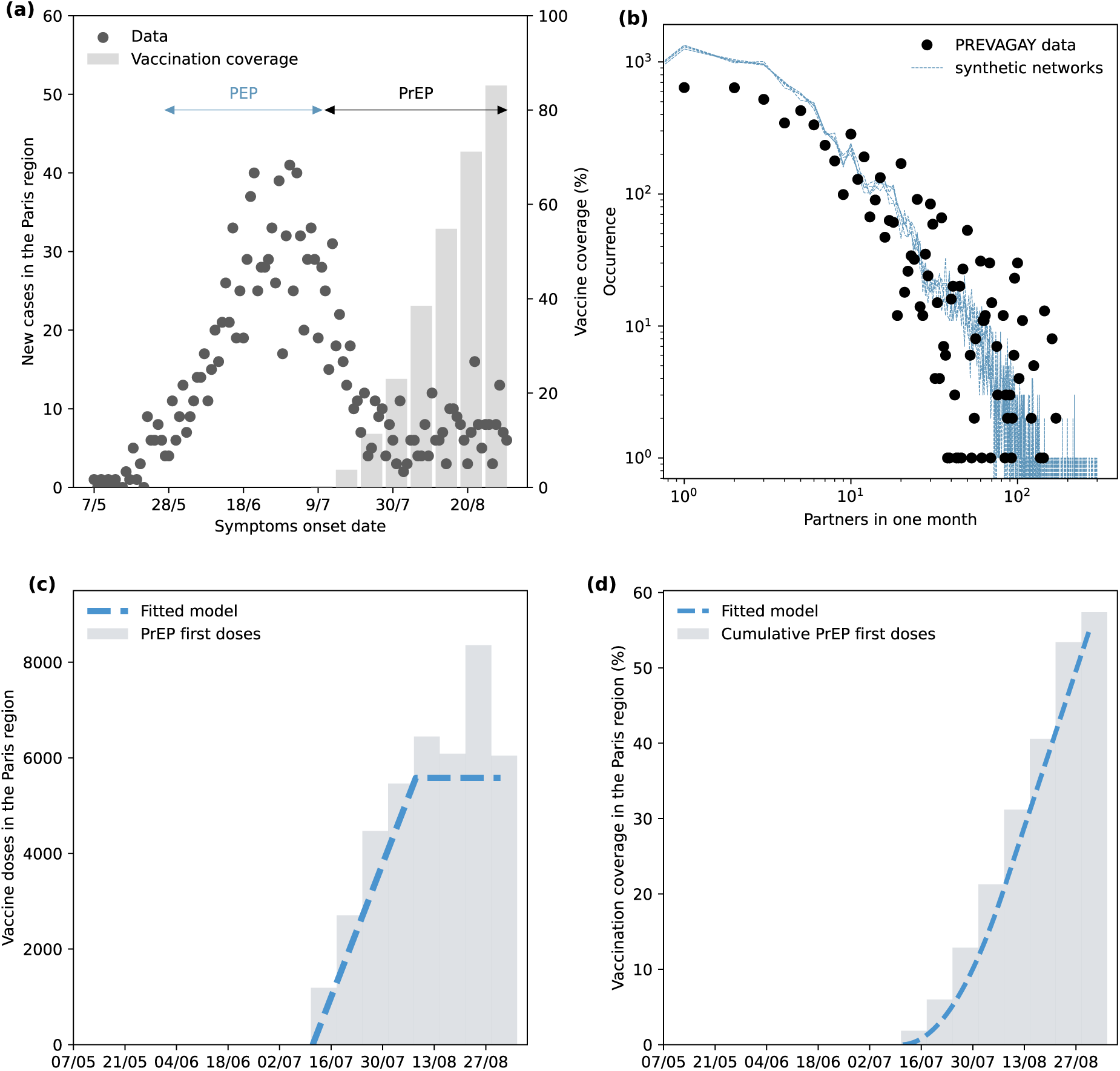
Surveillance, behavioral and vaccination data. **a:** Mpox cases in the Paris region from May 7 to August 31, 2022 (points, left y axis), together with 1^st^ vaccination “PrEP” (solid bars) in the Paris region (right y axis). The “PEP” and “PrEP” arrows refer to the periods when prophylaxis vaccination for cases’ contacts and pre-exposure vaccination were administered respectively. **b:** Occurrence of the monthly number of sexual partners in the PREVAGAY survey (points) and in the five synthetic networks (lines), in log-log scale. **c:** Vaccine 1^st^ doses (bars) in the Paris region and fitted functional form for the empirical doses distribution (dashed line). **d:** Vaccine 1^st^ doses vaccination coverage in the data (bars) and in the fitted model (dashed line).

### Mpox surveillance data

Data on the mpox outbreak were reported to Santé Publique France^21^ (French Public health Agency), including age, area of residence, dates of symptoms onset and testing, smallpox vaccination status and self-identification as a MSM (91% of answering cases). We accessed surveillance data for the Paris region, which included 1,616 mpox cases with symptoms onset or testing dates between May 7 and September 22, 2022. Among these, 33 cases had missing or misreported symptoms onset dates, but valid testing dates. For these, we considered symptoms onset already induced from the onset-to-testing delay obtained from cases with complete data (**Table S2**, **Figure S2**)^7^. During the study period from May 7 to August 31, 1,530 cases were included in the analysis, 30 of which with imputed onset dates.

### Vaccination data

Our model accounted for two types of vaccination: historical smallpox vaccination and the 2022 mpox vaccination campaign.

Smallpox vaccination in France was mandatory until 1979. Therefore, all MSM in the synthetic network born up to 1979 (47% of the population) were considered vaccinated against smallpox. We considered a vaccine effectiveness of 71%, estimated for France^22^.

During the 2022 outbreak, the third-generation Modified Vaccinia Ankara (MVA) smallpox vaccine Imvanex (called Jynneos in the USA) was used against mpox in France^23^. Vaccines became available on a voluntary basis starting July 11^24^. We accessed Santé Publique France data on the weekly number of administered doses in the Paris region (**Figure 1c, 1d**). We modeled the daily percentage of MSM to vaccinate with a ramp function, growing for the first four weeks before saturating until the end of our study period (**Figure 1c**). We refer to the slope of the linear growth of the ramp as the rollout speed *v*. We estimated the observed rollout speed *v\** = 0.05% fitting a linear model to the percentage of vaccinated MSM in the Paris region in the first four weeks from July 11, computed by dividing the administered vaccine doses for our population estimate (65,000 MSM) (**Figure S3**). We rescaled the resulting empirical weekly rollout speed to obtain the daily one (see details in the Supplementary Information). Depending on the scenario, we simulated mpox outbreaks under the observed rollout speed or we estimated it by fitting the model to the epidemic curve. We considered only MSM who had not yet been detected (i.e., those in the S, E, I, Id, and R compartments) eligible for vaccination. MSM already vaccinated against smallpox before 1979 were also eligible for vaccination. Vaccine was estimated to be effective only 14 days after administration with 78% effectiveness, as per available estimates^25,26^. A 74% effectiveness^27^, from more recent evidence, was also considered for sensitivity. In all scenarios, we tested two vaccination protocols, the first one where doses are administered irrespectively to sexual activity, and a second one where vaccine doses are administered to high-risk MSM preferentially. In this second case, we selected MSM to be vaccinated with a probability proportional to their number of monthly sexual partners.

In 2022, French public health authorities prioritized the broad distribution of first doses over the administration of second doses. Second-dose administration began only during the week of September 5–12, eight weeks after the vaccination campaign started and well after the epidemic peak in the Paris region (June 27–July 3). To reproduce this prioritization strategy, we excluded second doses from our main analysis but assessed their impact in the sensitivity analyses.

Also, some vaccine doses were available from May 27^23^ but in very limited supply and their use was restricted to the post-exposure prophylaxis for at-risk contacts of detected cases. We did not include the post-exposure prophylaxis vaccination in our model as it was incoherent with our study design, which considers preparedness scenarios where large vaccine stocks are available and used to initiate a pre-exposure vaccination campaign. Previous work provided evidence of the post-exposure prophylaxis vaccination not contributing to the mpox outbreak downturn in the Paris region^7^.

### PREVAGAY survey, time-varying sexual contact network construction, and MSM population estimate

Sexual contacts were represented through dynamic partnership networks parameterized using behavioral data from behavioral survey, capturing the age distribution, number of sexual partners and attendance at different types of sex-related venues^28,29^. We used data from the 2015 PREVAGAY survey of sexual behavior and HIV serostatus that was conducted among MSM in France^28,29^. The survey was designed to include MSM only, where MSM were self-declared. MSM were eligible if they were at least 18 years old, had sex with men in the previous 12 months, and could read and speak French. A total of 2,646 MSM reported socio-demographic data, such as age and place of residence, and sexual habits, in terms of number of sexual partners and attendance of MSM commercial venues. We used data from the 1,089 MSM living in Paris. 44% of these respondents, declared having more than 10 sexual partners in the previous month, and 8% declared more than 50 partners. Survey participants were asked to indicate if they attended gay venues in the past 12 months, including bars/clubs without sex, saunas with sex, backrooms/darkrooms, clubs/sex clubs and if they used internet dating sites or geolocalized gay dating apps.

From PREVAGAY data, we built stochastic time-varying networks of sexual contacts among MSM in the Paris region^28^. In this approach, MSM are represented by nodes in the network and time-varying sexual partnerships are represented by edges connecting two MSM and changing every day. Data allowed to describe sexual contacts occurring in specific venue type (saunas, backrooms, clubs, online dating) that we combined in a single network. We simulated partnerships in a synthetic population of 10,000 MSM, assigning to each MSM the following features taken from the PREVAGAY data according to sampling weights: age, monthly number of sexual partners, and information on his attendance or not to each venue type. We interpreted sexual contacts reported in the survey as single-day partnerships in our daily sexual contact networks. For each venue type, network realizations were generated by sampling from power-law distributions fitted to the venue-specific reported number of monthly sexual partners. The distribution of the monthly number of sexual contacts of the generated networks is shown in **Figure 1b**, and in **Figure S4** for the venue-specific plots. The fit is poorer when stratifying per venue, and this is likely influenced by rounding in self-reported partner numbers. Specifically, the network was constructed by assigning a number of monthly partners to each MSM for each venue type by sampling the corresponding fitted power law distribution six times, representing six months. To preserve the individual sexual behavior over time, the sampled numbers were sorted and assigned to individuals in the same order each month. We distributed the partners over time according to a multinomial distribution over the days of each month. We then generated the time-varying network corresponding to the sequence of 180 daily occurring partnerships in the MSM population using the configuration model^30^, pairing MSMs according to date and place of encounter^28,31^. The networks incorporated venue-based correlation, as individuals could only be connected if they reported attending the same type of venue. To take into account variability in the network building, we considered 5 stochastic realizations of the sexual network. For sensitivity, we also generated networks imposing a target power-law distribution with only 5% or 10% of MSM having more than 10 monthly partners, to evaluate epidemic outcomes in a population having a smaller fraction of high-risk individuals compared to the population under study (see details in the Supplementary Information).

The synthetic population of 10,000 MSM is a scaled representation of the estimated 65,000 MSM in the Paris region (26% of the estimated 250,000 multi-partner MSM in France^24,32^), used for numerical purposes. To compare simulation outputs to observed data—such as reported case counts or vaccine doses administered— we rescaled model outputs by a factor of 6.5 to align with the total target population size. In sensitivity analyses we varied MSM population size by ±20% to assess the impact of possible uncertainty on the estimate of the population size.

### Mpox transmission model

We developed a stochastic, data-driven network model of mpox transmission in the network of MSM in the Paris region parametrized as explained above. We described mpox transmission on the network of sexual contacts among MSM using a stochastic Susceptible-Exposed-Infected-Isolated-Recovered (SEIQR) agent-based model (**Figure S1**), adapted to include case detection. A susceptible individual (S) establishing a sexual contact with an infectious individual can contract the infection with a given probability (transmissibility *β)* per sexual contact. Following infection, the individual becomes exposed (E) but cannot yet transmit. After the latency period, he becomes infectious entering one of the two following infectious compartments: I_d_ if detected with detection probability *p_d_* = 60%, derived from previous work^7^, or I (undetected) otherwise. After the onset-to-testing period, I_d_ enters isolation (Q) where we assumed he abstains from any sexual contact, preventing therefore further transmission. After the infectious period or the isolation phase, the individual enters the recovery state (R, R_d_), becoming fully immune to the disease. The model was stratified to account for smallpox vaccination status at the start of the simulations, and rolling vaccination over time. The model was informed with the parameter values reported in **Table S1**; transmissibility per contact *β*and the date of mpox introduction were taken from our previous calibration to the 2022 epidemic^7^.

### Preparedness and community response scenarios

We evaluated four counterfactual scenarios combining different vaccination timings and rollout speeds with either community-wide reductions in sexual contacts or short-term post-vaccination abstinence. These scenarios tested whether earlier and faster pre-exposure vaccination could have reproduced the observed epidemic decline while eliminating or reducing the need for large-scale behavioral adaptation. All counterfactual scenarios assumed anticipated starts of the vaccination campaign with full access to vaccine. We explored four dates for the campaign start: May 20 (the day after the first case detection in France); May 27 (starting date of the public health response against mpox in the Paris region); June 3 and June 10, assuming a delay of 1 and 2 weeks with respect to May 27, respectively. For sensitivity, we explored May 7 – the day of the first alert of mpox in Europe^33^ – as the earliest possible date (**Figure S6**). In all scenarios, we compared a uniform vaccination campaign with one that prioritizes highly active MSM, with vaccination probability proportional to their number of sexual partners. We tested for sensitivity two additional vaccination protocols: a pragmatic one, where vaccine is given only to MSM with at least 50 or 100 monthly partners, and one where high-risk MSM are instead more reluctant to be vaccinated, resulting in low-risk MSM being preferentially vaccinated.

Scenarios differ in the assumptions on vaccination rollout and behavioral adaptations.

- **Earlier vaccination alone at observed rollout speed.** We simulated the outbreak with vaccination campaigns unfolding at the empirical speed *v\**. No behavioral adaptation was considered in this scenario. The resulting trajectory was then compared to the observed epidemic wave.
- **Earlier vaccination at observed rollout speed with behavioral change.** We simulated the outbreak with vaccination campaigns unfolding at the empirical pace *v*\* and incorporated behavioral adaptations. In our previous work^7^, both the magnitude of contact reduction *r* among MSM changing behavior and the overall fraction *c_r_* of MSM adopting this change were estimated by fitting the model to the 2022 epidemic curve, yielding a 90% reduction in sexual contacts among those adapting their behavior^7^. Here, we fixed *r* at this previously estimated value (*r* = 90%) and re-estimated *c_r_* for each vaccination start date, allowing the extent of behavioral adaptation required to vary with the contribution of vaccination to epidemic decline. In other words, we assumed that individuals adopting behavioral change reduced their sexual contacts by the same magnitude as estimated previously, while allowing the proportion of individuals adopting this change to vary according to the vaccination rollout. Behavioral change was introduced gradually from June 15 to July 21, as in previous work^7^. MSM were assumed to adopt behavioral changes with uniform probability, as this assumption provided the best fit to observations^7^. For sensitivity, we considered a scenario where we assumed only unvaccinated MSM as eligible for behavioral change and one in which higher-risk MSM preferentially changed behavior.
- **Earlier vaccination alone at accelerated rollout speed.** We simulated the outbreak varying the vaccine rollout speed, without any behavioral adaptations. We estimated the rollout speed *v* by fitting the model to the epidemic curve.
- **Earlier vaccination with accelerated rollout speed and post-vaccination abstinence.** We simulated earlier vaccination campaigns allowing the rollout speed *v* to vary and estimated the rollout speed required to reproduce the observed epidemic curve, as in the previous scenario. We additionally assumed that *c_a_* = 75% of vaccinated MSM complied with the recommendation of interrupting all sexual contacts during the 14 days following vaccine administration, corresponding to the period required to develop vaccine-induced protection, after which they returned to their usual sexual behavior. In sensitivity analyses, we considered 50% and 100% adherence to post-vaccination abstinence, as well as shorter durations (7 or 10 days) of the post-vaccination sexual abstinence period.

Scenario features and parameter values are summarized in **Table 1**.

**Table 1.** Scenario-specific features and parameter values.

| Scenario | Vaccination campaign start | Fitted parameters | Parameters inherited from previous work | Assumed parameters |
| --- | --- | --- | --- | --- |
| Earlier vaccination alone at observed rollout speed | May 20, 27;<br>June 3, 10 | - | $v^*=0.05\%$;<br>Date of mpox introduction=May 6;<br>$\beta = 0.29$ | - |
| Earlier vaccination at observed rollout speed with behavioral change | May 20, 27;<br>June 3, 10 | $c_r$ | $r=90\%$; behavioral changes period: from June 15 to July 20;<br>$v^*=0.05\%$;<br>Date of mpox introduction=May 6;<br>$\beta = 0.29$ | - |
| Earlier vaccination alone at accelerated rollout speed | May 20, 27;<br>June 3, 10 | $v$ | Date of mpox introduction=May 6;<br>$\beta = 0.29$ | - |
| Earlier vaccination with accelerated rollout speed and post-vaccination abstinence | May 20, 27;<br>June 3, 10 | $v$ | Date of mpox introduction=May 6;<br>$\beta = 0.29$ | $c_a=75\%$;<br>Duration of post-vaccination sexual abstinence =14 days |

### Inference framework

We simulated a population of 10,000 MSM and rescaled it to match the Paris region population (65,000). We averaged over 250 stochastic simulations to compute the expected epidemic curve and fitted the model to observed case counts through a (pseudo-) likelihood approach^34^.

The algorithm for the likelihood estimation works as follows: 1) first obtains the mean incidence over time by averaging 250 trajectories simulated with the model; then 2) uses the Poisson distribution to compute the likelihood. As we had at most one free parameter, we performed a grid search to find the maximum likelihood, using a quadratic approximation to smooth the response surface and locate the maximum. More details are provided in the Supplementary Information.

For the *Earlier vaccination at observed rollout speed with behavioral change* scenario, we fitted the model to the full epidemic curve and estimated *c_r_* (see above), associating 95% bootstrap confidence interval to its estimate. Also for the *Earlier vaccination alone at accelerated rollout speed* and the *Earlier vaccination with accelerated rollout speed and post-vaccination abstinence* scenarios we fitted the model to the full epidemic curve, this time to estimate *v* (see above), with 95% bootstrap confidence interval.

### Sensitivity analysis

We conducted eight additional analyses related to the *Earlier vaccination with accelerated rollout speed and post-vaccination abstinence* scenario. For all these additional tests, as in the main scenario, we estimated the rollout speed *v* fitting the model to the surveillance data.

- MSM population size. We assessed the robustness of our results to uncertainty in the estimated MSM population size by varying it by ±20%.
- Lower proportion of highly active MSM in the population. We simulated epidemic spread in networks where only 5% or 10% of MSM had at least 10 sexual partners per month, to assess the generalizability of our findings across MSM populations with different levels of sexual activity.
- Vaccination to low-risk MSM preferentially. We tested the hypothesis of the vaccination being distributed to MSM with a probability inversely proportional to the number of monthly partners
- Vaccination to MSM with at least 50 or 100 monthly partners. We tested the case where vaccination is administered only to MSM with more than 50 or 100 monthly partners.
- Vaccine effectiveness. We considered a vaccine effectiveness of 74%, estimated in a more recent statistical analysis comparing several estimates^27^.
- Second dose administration. We tested the impact of administering second doses, hypothesizing that 71% of the MSM who received a first dose would receive the booster, exactly four weeks later. 71% corresponds to the estimated percentage of first-dose recipients who received also a second dose between May 2022 and August 2024^35^.
- Adherence to post-vaccination sexual abstention. We tested the cases where 100% and 50% of the vaccinated MSM engage in post-vaccination sexual abstention
- Shorter duration of short-term post-vaccination sexual abstention. We hypothesized vaccinated MSM to engage in sexual abstention after vaccination for only 7 or 10 days, instead of 14 days.

### Ethics statement

Individual-level data were anonymized before use. Surveillance was considered as non-interventional research only requiring the non-opposition of the patient (article L1211 of the French public health code). The PREVAGAY study was authorized by the Comité de protection des personnes Île-de-France IX (n°2014-A01605– 42). Participants in the study gave their informed consent. Individual-level data were directly accessible to AM, HN, AT, AV in their capacity as employees of Santé publique France, the French national public health agency responsible for their collection and management. The other authors had access only to anonymized individual-level data, following authorization by Santé publique France through its data access procedure; no directly identifiable surveillance data were made available to them.

## Results

### Earlier vaccination alone at observed rollout speed

We first evaluated whether advancing the start of campaign, while maintaining the observed vaccination rollout rate observed in 2022, could have reproduced the epidemic decline in the absence of behavioral adaptation. When vaccination was distributed uniformly across MSM and initiated on May 20 – the day following the first detected case in France – the model led to a larger epidemic curve than observed (**Figure 2a**). The predicted incidence peak was delayed by 20 days (95%PI -13-51) and reached 2.1 times the observed magnitude (95%PI 0.0–4.5) (**Table 2**). By August 31, the model projected 2,922 cases (95%PI 29–5,335), nearly twice the number of cases observed in 2022. Thus, even under the earliest possible reactive campaign start, the observed rollout rate was insufficient to reproduce the observed decline. This limitation persisted under a more anticipatory scenario: starting vaccination on May 7, following the first alert of mpox in Europe^33^, still resulted in a larger and longer epidemic, with 2,330 projected cases (95%PI 54–4,319) (**Figure S7, Table S3**). Directing vaccination preferentially towards MSM with higher self-reported sexual activity substantially reduced epidemic size when the campaign was initiated early. With a May 20 start, prioritized vaccination resulted in 1,658 (95%PI 51–4,256) cases by August 31, only 9% more than reported cases, and producing an epidemic trajectory close to that observed (**Figure 2a**). Delaying the campaign start toward the empirical July 11 date in absence of behavioral change led to progressively larger epidemic waves and later declines (**Figure 2b–e**). This resulted in up to 220% (95%CI -95-360%) more cases than observed (**Figure 2f**), while the advantage of prioritizing high-risk MSM vaccination progressively diminished.

**Figure 2.**
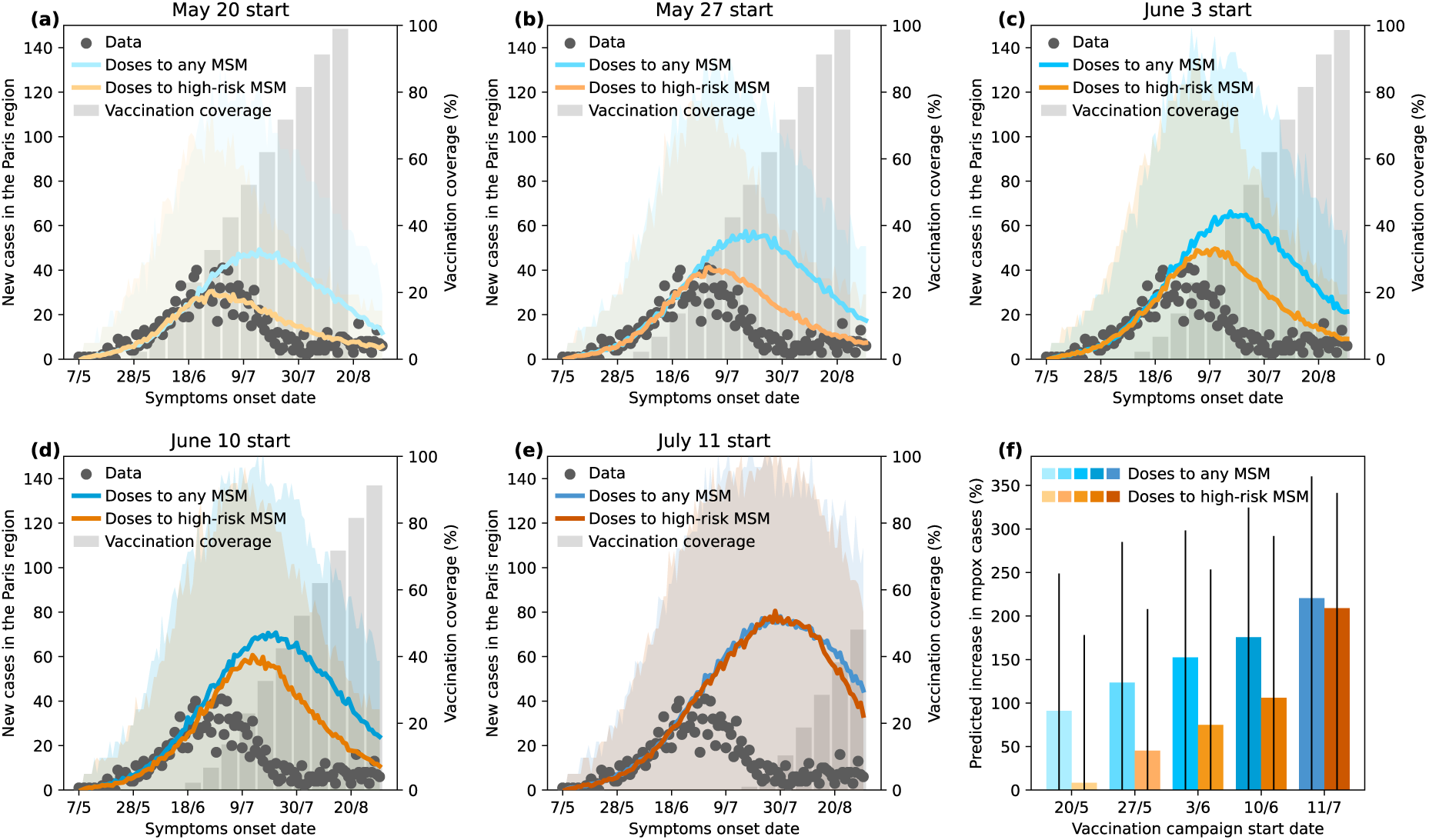
Model predictions for the *Earlier vaccination alone at the observed rollout speed* scenario. **a-e:** Mpox cases in the Paris region (points), mean model predictions across n = 250 stochastic simulations (lines), 95% prediction intervals (shaded areas), and vaccination coverage (bars). Each panel corresponds to a different vaccination campaign start date. **f:** Mean predicted increase in mpox cases relative to the observed cumulative number of cases through August 31, calculated across n = 250 stochastic simulations (bars), with 95% prediction intervals (lines)

**Table 2.** Scenario results.

| Scenario | Vaccination campaign type | Vaccination campaign start | Rollout speed | Modeled/ observed rollout speed | Weeks to reach 50% coverage | MSM reducing their sexual contacts | Cases at 31/8 | Cases at peak |
| --- | --- | --- | --- | --- | --- | --- | --- | --- |
| Earlier vaccination alone at observed rollout speed | Uniform | May 20 | Observed (0.050%) | 1 | 7 | No | 2,922 (29-5,335) | 66 (0-138) |
|  |  | May 27 |  |  |  |  | 3,421 (38-5,889) | 75 (0-144) |
|  |  | June 3 |  |  |  |  | 3,864 (74-6,093) | 83 (0-152) |
|  |  | June 10 |  |  |  |  | 4,217 (61-6,495) | 91 (0-153) |
|  |  | July 11 |  |  |  |  | 4,902 (70-7,044) | 108 (0-167) |
|  | High-risk MSM prioritized | May 20 |  |  |  |  | 4,902 (70-7,044) | 108 (0-167) |
|  |  | May 27 |  |  |  |  | 1,658 (51-4,256) | 38 (0-109) |
|  |  | June 3 |  |  |  |  | 2,221 (57-4,710) | 51 (0-123) |
|  |  | June 10 |  |  |  |  | 2,675 (55-5,407) | 61 (0-136) |
|  |  | July 11 |  |  |  |  | 4,727 (118-6,752) | 100 (0-165) |
| Earlier vaccination at observed rollout speed with behavioral change | Uniform | May 20 | Observed (0.050%) | 1 | 7 | 50.04% (47.73-52.36%) | 1,601 (32-3,492) | 42 (0-102) |
|  |  | May 27 |  |  |  | 55.22% (52.98-57.45%) | 1,490 (36-3,598) | 40 (0-100) |
|  |  | June 3 |  |  |  | 58.59% (56.34-60.83%) | 1,584 (29-3,801) | 42 (0-123) |
|  |  | June 10 |  |  |  | 60.21% (57.90-62.52%) | 1,694 (96-3,667) | 44 (0-109) |
|  |  | July 11 |  |  |  | 65.11% (63.10-67.11%) | 1,631 (44-3,787) | 42 (0-109) |
|  | High-risk MSM prioritized | May 20 |  |  |  | 16.28% (13.29-19.27%) | 1,444 (22-3,538) | 35 (0-87) |
|  |  | May 27 |  |  |  | 30.93% (27.46-34.39%) | 1,640 (23-4,053) | 44 (0-122) |
|  |  | June 3 |  |  |  | 42.80% (39.91-45.68%) | 1,580 (29-3,911) | 42(0-109) |
|  |  | June 10 |  |  |  | 52.82% (50.18-55.47%) | 1,546 (16-3,823) | 41 (0-115) |
|  |  | July 11 |  |  |  | 64.84% (62.88-66.81%) | 1,627 (29-3,669) | 42 (0-109) |
| Earlier vaccination alone at accelerated rollout speed | Uniform | May 20 | 0.22% (0.20-0.24%) | 4 (4-5) | 3.0 (2.9-3.1) | No | 1,525 (25-3,750) | 37 (0-100) |
|  |  | May 27 | 0.48% (0.44 - 0.53%) | 10 (9-11) | 2.0 (1.9-2.1) |  | 1,615 (44-3,769) | 39 (0-102) |
|  |  | June 3 | 3.05% (2.55 - 3.55%) | 51 (61-71) | 0.7 (0.7-0.9) |  | 1,348 (29-3,358) | 34 (0-100) |
|  |  | June 10 | 60.9% (52.3 - 69.5%) | 1,218 (1,046-1,390) | 0.1 (0.1-0.1) |  | 1,624 (32-3,668) | 39 (0-93) |
|  | High-risk MSM prioritized | May 20 | 0.08% (0.08-0.09%) | 1.6 (1.6-1.8) | 5.1 (4.7-5.1) |  | 1,414 (38-3,733) | 34 (0-94) |
|  |  | May 27 | 0.23% (0.21 - 0.24%) | 4.6 (4.2-4.8) | 2.9 (2.9-3.0) |  | 1,389 (23-3,739) | 34 (0-102) |
|  |  | June 3 | 0.51% (0.46 - 0.57%) | 10.2 (9.2-11.4) | 2.0 (1.9-2.0) |  | 1,562 (23-3,781) | 40 (0-115) |
|  |  | June 10 | 7.17% (6.17 - 8.17%) | 143.4 (123.4-163.4) | 0.4 (0.4-0.6) |  | 1,705 (72-3,984) | 44 (0-116) |
| Earlier vaccination with accelerated rollout speed and post-vaccination abstinence | Uniform | May 20 | 0.067 (0.065-0.07) | 1.34 (1.30-1.40) | 5.7 (5.6-5.9) | No – but 75% of the vaccinated MSM abstain from sex during the 14 days after vaccination | 1,276 (22-2,978) | 31 (0-80) |
|  |  | May 27 | 0.087 (0.083-0.09) | 1.74 (1.66-1.80) | 4.9 (4.7-5.0) |  | 1,240 (23-3,097) | 32 (0-94) |
|  |  | June 3 | 0.120 (0.114-0.126) | 2.40 (2.38-2.52) | 4.0 (4.0-4.1) |  | 1,559 (61-3,351) | 41 (0-102) |
|  |  | June 10 | 0.184 (0.172-0.197) | 3.68 (3.44-3.94) | 3.3 (3.1-3.4) |  | 1,455 (23-3,478) | 40 (0-109) |
|  | High-risk MSM prioritized | May 20 | 0.017 (0.016-0.018) | 0.34 (0.32-0.36) | 17.0 (16.1-17.9) |  | 1,295 (31-3,387) | 26 (0-73) |
|  |  | May 27 | 0.024 (0.022-0.025) | 0.48 (0.44-0.50) | 12.6 (12.1-13.6) |  | 1,352 (14-3,465) | 28 (0-80) |
|  |  | June 3 | 0.035 (0.033-0.037) | 0.70 (0.66-0.74) | 9.3 (8.9-9.7) |  | 1,344 (25-3,487) | 30 (0-94) |
|  |  | June 10 | 0.061 (0.055-0.067) | 1.22 (1.10-1.34) | 6.1 (5.7-6.6) |  | 1,326 (22-3,279) | 33 (0-101) |

### Earlier vaccination at observed rollout speed with behavioral change

Since earlier vaccination at the observed rollout rate did not suffice to reproduce the observed epidemic decline, we next quantified how much behavioral adaptation would have been required given earlier campaign initiation (**Figure 3**). For a May 20 start, 50.0% (95%CI 47.7–52.4%) of the Paris MSM population would need to substantially reduce their sexual contacts (**Figure 3a, f**), compared to 65.1% (95%CI 63.1–67.1%) under the empirical July 11 start (**Figure 3e, f; Table 2**). This corresponds to a 23% relative reduction in the proportion of MSM needing to change behavior. This estimate assumes that individuals adopting behavioral change reduce their sexual contacts by 90%, as estimated in our previous work^7^, with this magnitude of individual-level contact reduction held constant across vaccination start dates. Even with an earlier start on May 7, more than one third of MSM would still need to substantially reduce their risk (**Figure S8**, **Table S3**), indicating that earlier vaccination at the observed rollout rate would have reduced – but not eliminated – the need for large-scale behavioral adaptation. Sensitivity analyses showed that results were robust when behavioral adaptation were limited to unvaccinated MSM and that concentrating behavioral change among highly sexually active MSM substantially reduced the proportion of the community required to change behavior (**Figures S11, S12; Tables S4, S5**).

**Figure 3.**
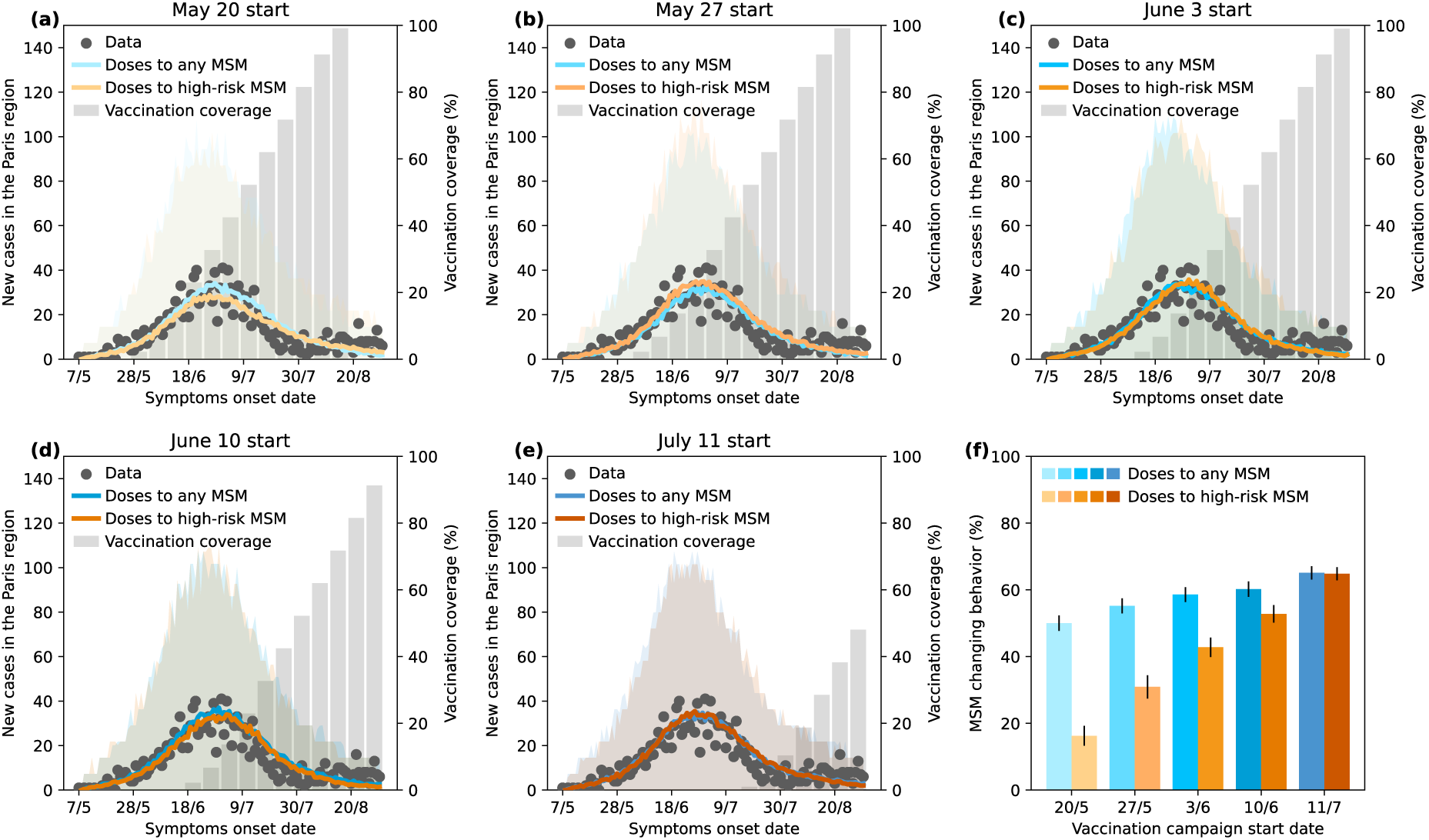
Model predictions for *the Earlier vaccination at observed rollout speed with behavioral change* scenario. **a-e:** Mpox cases in the Paris region (points), mean model predictions across n=250 stochastic simulations (lines), 95% prediction intervals (shaded areas), and vaccination coverage (bars). Each panel corresponds to a different vaccination campaign start date; panel e corresponds to July 11, the empirical start date of the 2022 pre-exposure vaccination campaign, and corresponds to the scenario considered in previous work. **f:** Percentage of MSM changing behavior: maximum-likelihood estimates (bars), with 95% confidence intervals (lines).

Whereas uniform vaccination only moderately reduced the need for behavioral adaptation (23% relative reduction), prioritizing vaccination to highly sexually active MSM resulted in a substantially lower proportion of the community needing to change behavior (16.3% (95% CI 13.3–19.3%)) for a May 20 start. This is to be compared to 64.8% (95% CI 62.9–66.8%) for a July 11 start, corresponding to 75% relative reduction; **Figure 3f**).

### Earlier vaccination alone at accelerated rollout speed

We next assessed whether accelerating the vaccination rollout – as well as advancing the campaign start – could have limited transmission to the level observed in 2022 without behavioral adaptation (**Figure 4**). The required vaccination rollout speeds were substantially higher than the observed 2022 rate. For vaccines uniformly distributed among MSM, the required rollout speed was 4.4 (95%CI 4.0-4.8) times faster than the observed speed for a May 20 start and increased to an unrealistic 1,218 (95%CI 1,046-1,390) times faster for a June 10 start (**Figure 4e**, **Table 2**). Consequently, the time to reach 50% coverage shortened from 3 down to 0.1 weeks, instead of the observed 7 weeks (**Figure 4f**). Even with a May 7 start, the rollout speed would still need to more than double relative to the observed rate (**Figure S9, Table S3**). With prioritized vaccination, the required acceleration was 1.6 (95%PI 1.6-1.8) to 143 (95%CI 123-163) times the observed rate, with time to 50% coverage shortening from 5 down to 0.4 weeks.

**Figure 4.**
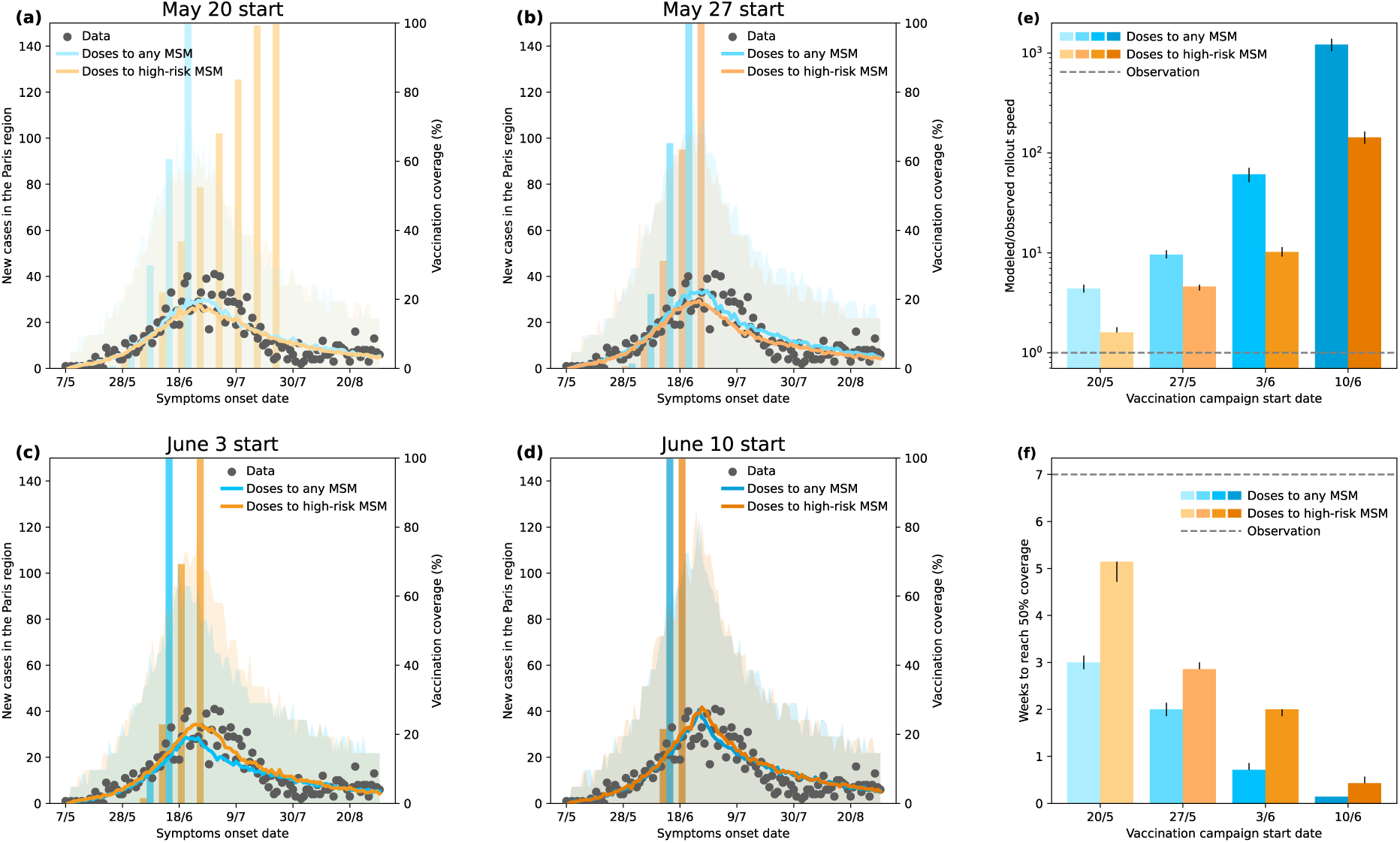
Model predictions for the *Earlier vaccination alone at accelerated rollout speed* scenario. **a-d:** Mpox cases in the Paris region (points), mean model predictions across n = 250 stochastic simulations (lines), 95% prediction intervals (shaded areas), and vaccination coverage (bars). Each panel corresponds to a different vaccination campaign start date. **e:** Ratios of the maximum-likelihood estimates of the model-predicted rollout speeds to the observed rollout speed (bars), with corresponding 95% confidence intervals (vertical lines). The dashed horizontal line (y=1) represents the reference for a predicted rollout speed equal to the observed one. **f:** Number of weeks needed to reach 50% vaccination coverage, derived from the maximum-likelihood estimates of the model predicted rollout speeds (bars), with corresponding 95% confidence intervals (vertical lines). The dashed horizontal line (y=7) represents the empirical number of weeks.

### Earlier vaccination with accelerated rollout speed and post-vaccination abstinence

To offset the substantial increase in vaccination rollout required above, we assessed whether combining faster rollout with short-term behavioral adaptation could reduce the overall vaccination effort. Specifically, we evaluated the effect of post-vaccination sexual abstinence during the 14-day period needed to develop vaccine-induced protection, assuming 75% compliance (**Figure 5**). The rollout speed required under uniform vaccination decreased markedly compared with the scenario without sexual abstinence. Depending on the campaign start date (May 20–June 10), the required rollout ranged from 1.34 (95%CI 1.30-1.40) to 3.68 (95%CI 3.44-3.94) times the empirical rate (**Figure 5e**, **Table 2**), i.e. 70% (95%CI 68-70%) to 99.7% (95%CI 99.7-99.7%) lower than the values without behavioral adaptation. This corresponded to reaching 50% vaccination coverage in 3.3–5.7 weeks, closer to the observed 7 weeks (**Figure 5f**, **Table 2**). Preferential vaccination of high-risk MSM further reduced the required acceleration, with rollout speeds ranging from 0.34 (95%CI 0.32-0.36) to 1.22 (95%CI 1.10-1.34) times the empirical rate. Post-vaccination abstinence would involve at most about 50% of MSM abstaining simultaneously when vaccination was distributed uniformly (**Figure 5g**), but substantially fewer when vaccination targeted high-risk MSM (**Figure 5g, h**). At the population level, the resulting proportion of daily sexual contacts averted peaked at 71% for a uniform vaccination campaign starting on June 10, while remaining below 50% across all campaigns targeting high-risk MSM (**Figures S5a, S5b**).

**Figure 5.**
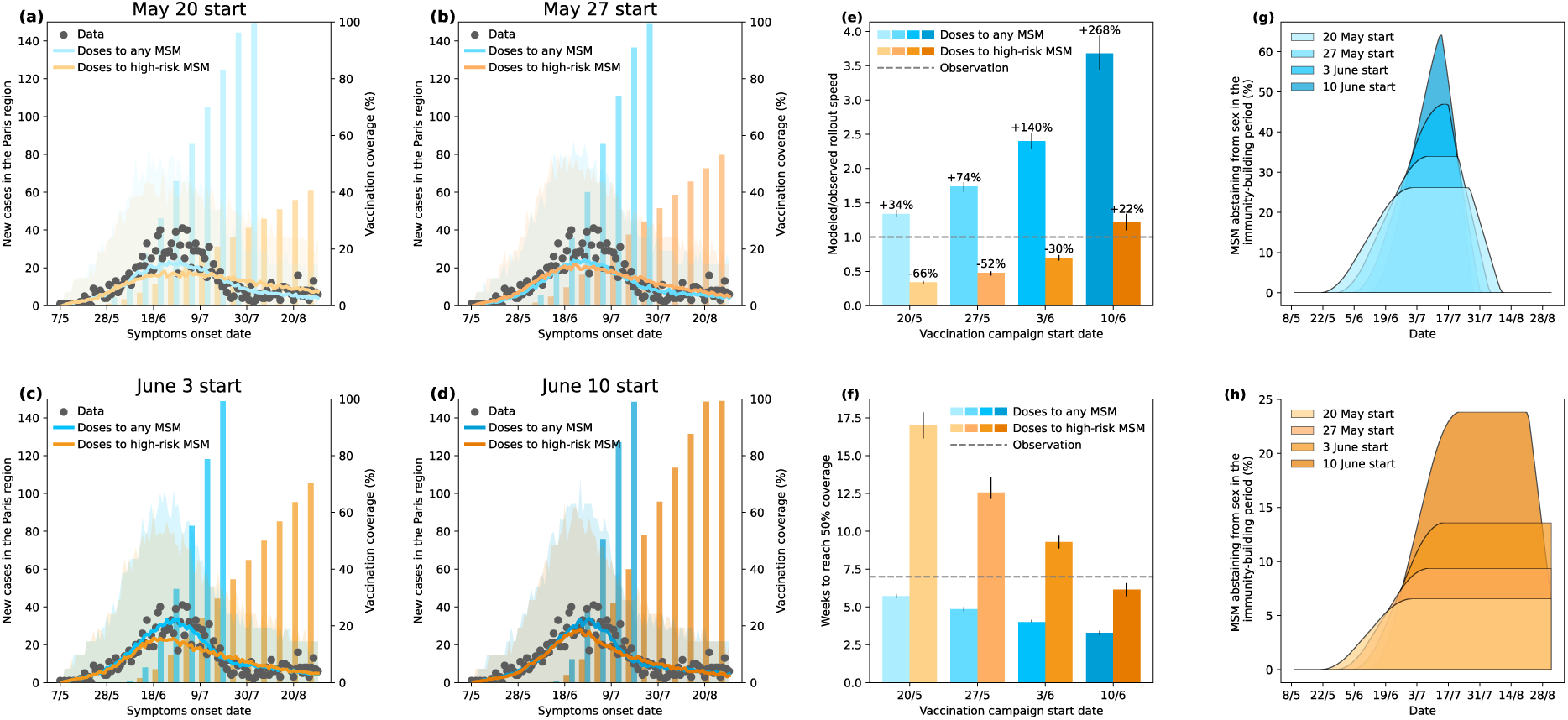
Model predictions for the *Earlier vaccination with accelerated rollout speed and post-vaccination abstinence* scenario. **a-d:** Mpox cases in the Paris region (points), mean model predictions across n = 250 stochastic simulations (lines), 95% prediction intervals (shaded areas), and vaccination coverage (bars). Each panel corresponds to a different vaccination campaign start date. **e:** Ratios of the maximum-likelihood estimates of the model-predicted rollout speeds to the observed rollout speed (bars), with corresponding 95% confidence intervals (vertical lines). The dashed horizontal line (y=1) represents the reference for a predicted rollout speed equal to the observed one. **f:** Number of weeks needed to reach 50% vaccination coverage, derived from the maximum-likelihood estimates of the model predicted rollout speeds (bars), with corresponding 95% confidence intervals (vertical lines). The dashed horizontal line (y=7) represents the empirical number of weeks. **g, h:** Mean daily percentage of MSM abstaining from sex due to post-vaccination sexual abstinence if vaccination is given to any MSM (g) or to high-risk MSM preferentially (h), calculated across n=250 simulations.

Sensitivity analyses confirmed these trends: with 50% compliance to 14-day sexual abstinence after vaccine administration, the required rollout speed increased to 1.68 (95%CI 1.60-1.76) to 7.14 (95%CI 6.48-7.78) times the empirical rate (still 62% (95%CI 60-64%) to 99.4% (95%CI 99.4-99.5%) lower than the values without behavioral adaptation), whereas 100% compliance reduced it to 1.12 (95%CI 1.08-1.16) to 2.60 (95%CI 2.48-2.72) times (still 75% (95%CI 74-75%) to 99.8% (95%CI 99.8-99.8%) lower), depending on the vaccination strategy and the campaign start date (**Figures S26–S28, Table S14**). The estimated rollout rate was lower than the observed one only if the campaign started on May 7 (**Figure S10, Table S3**).

To test the robustness of these findings, we examined alternative assumptions on vaccine effectiveness, vaccine uptake and prioritization, compliance to recommendations and MSM population size and structure. Assuming that highly sexually active MSM were more reluctant to be vaccinated—so that vaccination reached lower-risk individuals preferentially—led to rollout speeds very similar to the ones obtained under uniform vaccination (e.g., 1.28 (95%CI 1.22–1.32) times the observed rate for a May 20 start, compared with 1.34 under uniform vaccination (**Figure S20**, **Table S10**)). This is a consequence of the heavy-tailed structure of sexual contacts (**Figure 1b**): because most MSM have relatively few partners, uniform vaccination also predominantly reaches lower-risk individuals. As a more pragmatic prioritization strategy, we also tested vaccinating MSM above various thresholds of monthly sexual partners. Restricting vaccination to individuals with at least 100 partners per month consistently resulted in rollout speeds below the observed rate for all campaign start dates. Lowering the threshold to at least 50 partners altered this pattern, with rollout speeds exceeding the observed rate for later campaign start dates (**Figure S21**, **Table S11**). This reflects the benefit of rapidly immunizing the most highly connected individuals in heavy-tailed contact networks, who are reached early by the epidemic and contribute disproportionately to transmission. Higher rollout speeds were also required when considering shorter duration of post-vaccination sexual abstinence: they were estimated to be +147% (95%CI 125–170%), for a widespread campaign starting June 10 assuming 10 days of abstinence, and increased to +383% (95%CI 334–431%) assuming 7 days of abstinence (**Figures S29-S31, Table S15**), as vaccination compensated for increased opportunities of transmission. We also tested a vaccine effectiveness of 74%, drawn from a recent systematic search and meta-analysis of the available data^27^; as this value is close to the 78% used in our main analysis, results were largely unchanged (**Figures S22, S23; Table S12**). We further considered the inclusion of second-dose administration, which was excluded from the main analysis to reproduce the 2022 strategy of prioritizing broad first-dose coverage over booster administration. Including second doses did not substantially affect the results (**Figure S25; Table S13**): even for the May 20 and May 27 campaign starts, where second-dose administration began before the epidemic peak (**Figure S24**), the 14-day delay before vaccine effectiveness meant that its protective effect was only realized after the peak. We also tested populations with substantially smaller proportions of highly active MSM (5% or 10% with at least 10 monthly sexual partners). This had limited effects on vaccination requirements under widespread vaccination, while high-risk prioritization remained highly efficient and required even lower rollout speeds, as fewer doses were needed to reach the smaller group disproportionately contributing to transmission (**Figures S17–S19, Table S9**). Finally, we assessed sensitivity to uncertainty in the estimated MSM population size by varying it by ±20%. The rollout speeds required to reproduce the observed epidemic trajectory were only moderately affected, and the qualitative conclusions remained unchanged (**Figures S13–S15**). Time to 50% coverage was similarly robust under widespread vaccination (**Figure S15c**), whereas under high-risk-targeted vaccination it varied non-monotonically with population size (**Figure S15d**). This results from competing effects: a smaller population requires vaccinating fewer high-risk MSM to achieve a comparable epidemiological effect, allowing a slower rollout, while the opposite occurs in a larger population.

## Discussion

Using the 2022 mpox outbreak in the Paris region as an empirical benchmark, we show that attempting to control the epidemic by vaccination alone, under realistic rollout conditions, would have led to much larger number of cases. Even when initiated earlier, vaccination at the observed 2022 rollout speed only modestly reduced the need for behavioral adaptation, with large-scale reductions in sexual contacts still required. Achieving a comparable epidemic trajectory through vaccination alone would have required at least a 4-fold faster deployment, which we consider currently challenging. In contrast, combining targeted vaccination with short-term behavioral measures – such as temporary risk reduction during the period of vaccine-induced immunity building – substantially improved epidemic mitigation. These findings indicate that epidemic control in this context depends on the joint dynamics of vaccination and changes in human behavior. Preparedness and response strategies should explicitly account for their interaction rather than treating these as independent levels.

Advancing the start of vaccination alone is insufficient to substantially alter epidemic outcomes in the absence of adequate rollout capacity. Even under the earliest plausible reactive start, vaccination at the observed 2022 rollout speed would not have achieved a decline comparable to that observed and only moderately reduced the behavioral adaptation required. Instead, the dominant constraint was the speed at which vaccination could be delivered: achieving similar epidemic trajectories through vaccination alone would have required rollout rates several-fold higher than those observed, even under favorable conditions. Such a rollout speed was not observed in any European country; moreover, France already emerged in 2022 as the European country who administered the largest number of doses^36^. These findings suggest that preparedness efforts should prioritize not only early detection and rapid initiation of vaccination, but also the capacity to scale up delivery rapidly in the initial phase of the outbreak. In practice, this implies that investments in logistics, supply chains and access to vaccination services may be as critical as surveillance systems in determining the collective effectiveness of vaccination-based responses.

Our results further show that combining vaccination with short-term behavioral recommendations can substantially improve the feasibility of epidemic mitigation under realistic rollout constraints. The challenges associated with promoting sustained reductions in sexual activity among MSM may instead be addressed through temporary risk-reduction recommendations linked to vaccination, during the short period required to develop vaccine-induced protection. This approach markedly reduced the vaccination effort needed to achieve a decline comparable to that observed in 2022. Under moderate to high compliance (50% to 75%), the required rollout speed decreased to levels much closer to those observed (about 1.7 and 1.3 times faster, respectively), compared with substantially higher rates in the absence of behavioral adaptation (>4 times faster). Importantly, this response is temporally and socially localized, involving only a subset of individuals for a limited duration, rather than the large-scale, sustained reductions in contacts observed during the 2022 outbreak^7^. In practice, such recommendations could be delivered at the point of care, with clinicians advising temporary risk reduction while vaccine-induced protection develops. This approach may also enhance uptake, as healthcare professionals were identified in some settings as the most trusted information source among MSM during the 2022 outbreak^37^. However, reports of insufficient information^38^ and difficulties interacting with healthcare providers during the 2022 wave in France^39^, together with the central role of digital platforms and community associations in information dissemination^40^, suggest that complementary communication channels may be needed to effectively reach diverse at-risk populations. These considerations motivated our exploration of different compliance levels to capture plausible heterogeneity in behavioral adherence.

Targeted vaccination strategies are critical for improving epidemic control in heterogeneous contact networks. Consistent with theoretical expectations for heavy-tailed distributions of sexual partnerships^41–43^, prioritizing individuals with the highest levels of sexual activity substantially increased vaccination campaign effectiveness and reduced the rollout speed required to achieve a comparable epidemic decline. This reflects the rapid early spread of infection within highly active subgroups, which can sustain and amplify transmission^44–46^ if not reached early by vaccination. However, the benefits of prioritization depend critically on both timing and implementation. While preferential vaccination of highly active MSM yields large gains when implemented early, its advantage over uniform strategies diminishes rapidly as vaccination is delayed, because infection reaches high-risk individuals before vaccination. Moreover, more pragmatic prioritization approaches based on thresholds of partner numbers showed more variable outcomes and, in some cases, reduced efficiency gains. Although based on the same information, such threshold-based strategies treat all individuals above a cutoff similarly: lowering the threshold (e.g., from 100 to 50 partners) enlarges the target group but dilutes the focus on those at highest risk, particularly as roll-out delays increase. These findings highlight the challenges of translating theoretically optimal prioritization into practice, where both timely deployment and accurate identification of high-risk individuals are required. Encouragingly, evidence suggests that individuals at highest risk may also be more willing to vaccinate^47^, which facilitates targeted strategies. But in addition, real-world implementation is constrained by logistical factors affecting access and scale-up. Although willingness to vaccinate generally increases with perceived risk^18,48–50^, lack of access was a primary barrier during the 2022 outbreak, especially due to stock scarcity^51^. Effective prioritization depends not only on identifying high-risk individuals, but also on ensuring rapid access to vaccination, supported by sufficient infrastructure, workforce capacity, and community engagement^52^.

The central role of vaccination timing and rollout capacity identified here has direct implications for the current mpox epidemiological context, characterized by persistent low-level global circulation of clade IIb alongside the rapid emergence and geographic expansion of clade I lineages^1^. Despite the decline of the 2022 global outbreak, transmission has not been interrupted. Recurrent clusters and evolving transmission patterns are reported across regions, including Europe^1^. Meanwhile, accumulating immunological evidence indicates that population-level protection may be progressively eroding^53,54^. This waning immunity, combined with the growing cohort of individuals with no prior exposure to orthopoxviruses, creates conditions for renewed susceptibility and potential resurgence. Also, waning immunity may have the effect of reducing symptoms in infected individuals, provoking detection challenges^55^. Maintaining sufficient levels of protection in at-risk populations may also be critical to mitigate future resurgences.

This study presents several limitations. First, community-wide behavioral adaptations (not linked to vaccination) were modeled using simplified assumptions on a uniform magnitude of contact reduction, estimated in previous work^7^, without capturing possible heterogeneity across individuals and over time or dynamic feedback with perceived risk. Short-term behavioral changes after vaccination as well, in real-world, would be likely to vary across subgroups, reflecting differences in risk perception, integration into care, and exposure to tailored information. We explored different compliance levels (50–100%) to compensate for this aspect. Overall, behavioral responses are inherently difficult to predict and may vary across populations and outbreaks depending on risk perception, community engagement, public health messaging, and the epidemiological context. Our counterfactual estimates should therefore be interpreted as conditional on the behavioral response considered in each scenario, rather than as predictions of the behavioral response that would occur in a future outbreak. Second, our networks constructed from sexual behavioral survey data are not degree-assortative. However, in previous work^7^ we showed that sensitivity analyses increasing degree assortativity levels had no impact on the results. By contrast, venue-based correlation is intrinsic to the network construction—derived from co-attendance to specific venue types—and reflects the increased likelihood of connection among individuals sharing the same sexual and social contexts. Third, we did not model pre-symptomatic transmission^56^, similarly to other modeling studies^9,10,15^, as previous analyses show that this assumption does not alter epidemic outcomes^7^. Fourth, the analysis focused on the specific epidemiological and social context of the 2022 outbreak, when the population was largely immunologically naïve to mpox, which may limit direct quantitative generalization. Future outbreaks may occur in populations with different levels of vaccine-and infection-derived immunity, vaccine uptake, behavioral responses to a novel epidemic wave, potentially leading to different quantitative outcomes. Nevertheless, the mechanisms highlighted by our counterfactual analyses—particularly the interplay between vaccination timing, rollout capacity, prioritization, and behavioral responses—are expected to remain relevant across different epidemic contexts.

Mpox control under realistic conditions cannot rely on vaccination alone. Instead, epidemic outcomes depend on the combined effects of vaccination timing, rollout capacity and behavioral responses. Combining rapid vaccination with short-term behavioral measures – such as 14-day post-vaccination sexual abstinence recommended at the point of care – makes epidemic control achievable. Targeted information campaigns promoting vaccine uptake among MSM with higher numbers of sexual partners remain essential, as succeeding in reaching this population can substantially improve outbreak control. The recent identification of Clade Ib cases in Europe^6^ reinforces the continued risk of resurgence, alongside waning immunity^53,54^ and the progressive entry of younger, sexually active but immunologically naïve individuals into at-risk groups.

Designing preparedness strategies that align intervention capacity with transmission dynamics and health promotion will be essential to limit the impact of future mpox outbreaks and similar emerging infections.

## Data availability

Access to individual-level data from the PREVAGAY survey and surveillance data, which contain sensitive information on sexual behavior and health, is restricted. These data, alongside with vaccination data, which were not published, can be made available to researchers upon request to Annie Velter, subject to approval of the proposed analyses and agreement to adhere to security, confidentiality and collaborative conditions. The timeframe for access will depend on the review and approval process.

## Code availability

Our code, usable to reproduce all our experiments, is publicly available at the GitHub repository (https://github.com/EPIcx-lab/mpox-behavioral-2). Simulations were developed and executed in C++ (version 14.2.0). Data processing and analyses were carried out in Python (version 3.8.10). Network construction and the quadratic approximation of the fitting procedure were carried out using R (version 4.2.3).

## Authors’ contributions

VC conceived and designed the study. DM, OR, PYB contributed to the methodology. OR and PYB developed the code for the generation of the MSM sexual network. DM developed the code for the inference and the modeling of the transmission dynamics, ran the simulations, and analyzed the data. All authors interpreted the results. DM and VC wrote the initial manuscript draft. DM, OR, PYB, HN, AM, AT, AV, VC edited and approved the final version of the Article.

## Acknowledgements

We thank Anne-Sophie Barret, Emilie Chazelle, and Laura Zanetti for useful discussions.

## Footnotes

* Men-who-have-sex-with-men (MSM) refers to all men who engage in sexual relations with other men. The words “men” and “sex” are interpreted differently in diverse cultures and societies and by the individuals involved. Therefore, the term encompasses the large variety of settings and contexts in which male-to-male sex takes place, regardless of multiple motivations for engaging in sex, self-determined sexual and gender identities and various identifications with any particular community or social group (Consolidated guidelines on HIV, viral hepatitis and STI prevention, diagnosis, treatment and care for key populations. Geneva: World Health Organization; 2022 (https://apps.who.int/iris/handle/10665/360601, accessed September 1, 2023).).

